# The economic value of quarantine is higher at lower case prevalence, with quarantine justified at lower risk of infection

**DOI:** 10.1101/2020.11.24.20238204

**Authors:** James Petrie, Joanna Masel

**Affiliations:** Applied Mathematics, University of Waterloo; WeHealth Solutions PBC; Ecology & Evolutionary Biology, University of Arizona

## Abstract

Some infectious diseases, such as COVID-19, are so harmful that they justify broad scale social distancing. Targeted quarantine can reduce the amount of indiscriminate social distancing needed to control transmission. Finding the optimal balance between targeted vs. broad scale policies can be operationalized by minimizing the total amount of social isolation needed to achieve a target reproductive number. Optimality is achieved by quarantining on the basis of a risk threshold that depends strongly on current disease prevalence, suggesting that very different disease control policies should be used at different times or places. Aggressive quarantine is warranted given low disease prevalence, while populations with a higher base rate of infection should rely more on social distancing by all. The total value of a quarantine policy rises as case counts fall, is relatively insensitive to vaccination unless the vaccinated are exempt from distancing policies, and is substantially increased by the availability of modestly more information about individual risk of infectiousness.

## 2. Introduction

Control of SARS-CoV-2 transmission has not been achieved by interventions such as improved ventilation alone, but has also required social distancing. Social distancing can take the form either of population-wide measures, ranging from extreme, mandatory lockdowns to more modest voluntary behavior change, or it can be targeted via effective testing, tracing, quarantine, and isolation. Social distancing to control the spread of SARS-CoV-2 has imposed immense social costs, and is difficult to maintain for long periods. Targeting quarantine to those at higher risk of being infectious has the potential to reduce the total harms of social distancing across the population. Here we calculate which combination of population-wide social distancing plus targeted quarantine will minimize harms while controlling transmission. We operationalize this question by comparing the harms of different policy combinations that all achieve the same target value for the effective reproduction number. Figure 1 demonstrates how quarantining a greater fraction of infectious individuals allows overall social distancing to be reduced while holding the number of expected transmissions constant.

**Fig. 1.**
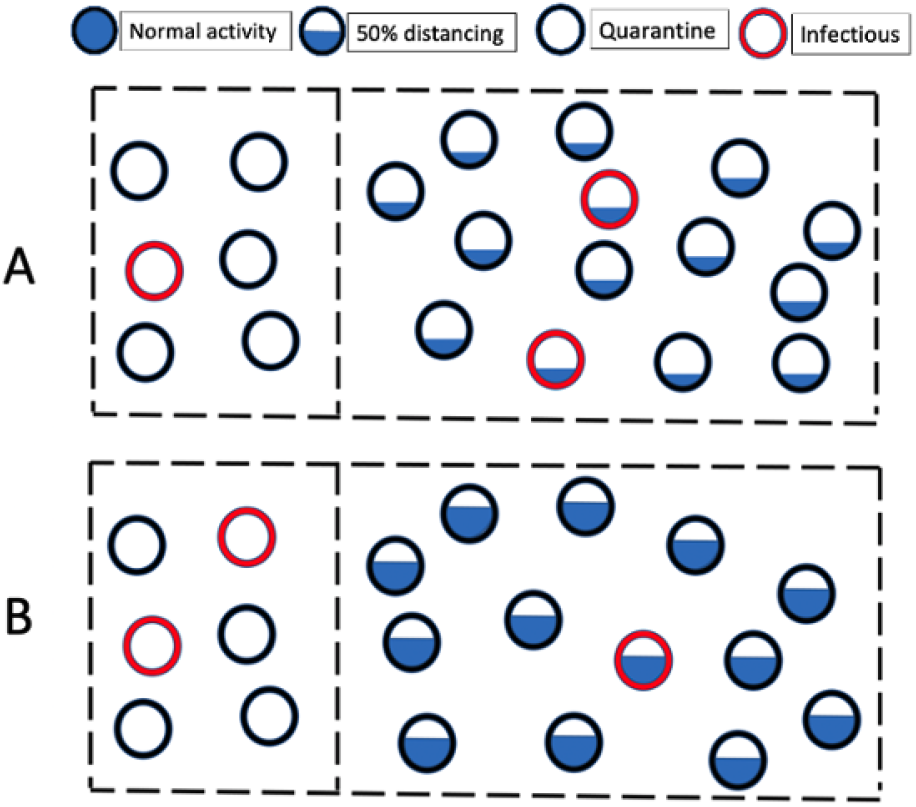
Two scenarios of quarantine and social distancing that give rise to the same number of expected transmissions. A) One out of three infectious individuals is quarantined, and the general population is at 1/3 of normal social activity. B) Two out of three infectious individuals are quarantined, and the general population is at 2/3 of normal social activity. By finding and quarantining infectious people, social distancing imposed on the general population is significantly reduced.

Our approach, at the interface of infectious disease epidemiology and economics (1), solves a number of problems previously encountered at that interface. First, it is difficult to decide on an exchange rate between social restrictions / livelihoods versus lives (2); compounding this is the fact that selfish agents may count only the cost to their own life, and not that of others they infect (3). Difficulties determining an exchange rate lead some to present only idealized curves without a concrete recommendation (4, 5). Idealized curves can lead to abstract insights, but are of limited use to practical decision-making.

We avoid the need for an exchange rate between social restrictions and infections by instead comparing one form of (population-wide, partial) social restrictions with another form of (targeted, more stringent) social restrictions. This allows the lowest cost strategy to be selected without having to monetarize the cost of health outcomes, the latter being held constant in our analysis. The policy optimization problem is thus framed as a minimization of a cost function across different degrees of isolation for different individuals.

Second, much prior work integrates across an entire sweep of a pandemic modelled using an SIR approach (6). These projections have often been inaccurate. We instead focus on a moment in time, an approach that is enabled by a control theory view. At the beginning of the COVID-19 pandemic, populations experienced rapid exponential growth, e.g. doubling in as little as 2.75 days in New York City (7). Through some combination of top-down control measures and individual behavior modifications, many populations subsequently achieved relatively flat case counts, i.e. an effective R value remarkably close to 1 (8–10). Even locations that subsequently lost control of the pandemic have tended to eventually issue stay at home orders leading to exponential decline. Over the very long term, a geometric mean of R not much greater than 1 is inevitable (see Appendix A). This implies that there is a form of control system (whether intentional or not) using social distancing to regulate transmission rate.

Third, many models consider decision-making that is informed by perfect, instantaneous knowledge (6). This is problematic given that key indicators such as hospital usage or even positive tests have a marked lag relative to infections. Our approach focuses instead on making optimal use of available, probabilistic information. Past economic approaches to modelling the transmission of SARS-CoV-2 treat more stark examples of information, e.g. testing to find out who is infected and should isolate, as opposed to who is exposed to what degree and should quarantine (6, 11). But quantitative information about individual risk can be obtained from a variety of sources. For example, the risk of infection with SARS-CoV-2 can be estimated based on proximity and duration of contact with a known case, and their estimated infectiousness as a function of timing relative to symptom onset date (12). Similarly, setting local rather than global shutdown policies, in the light of differences between regions, can lower costs (13).

Our work ultimately describes the value of information, specifically information about who is at high enough risk of being infected to quarantine strictly rather than merely to conform to population-wide restrictions. Given an individual estimated to be infectious with probability ‘r’, we propose a method for deciding whether to quarantine this individual. We do so by weighing the cost of quarantine against the degree to which indiscriminately applied social distancing would need to be increased to achieve the same reduction in transmission. Our approach informs choice of the lowest cost strategy needed to achieve epidemiological targets. In particular, we formalize the intuition that populations with low prevalence should widen the net of who they quarantine.

## 3. Model

### A. Optimal Risk Threshold

Consider a well-mixed population of size *P*, of which *I* people are currently both infected and infectious, and *S* people are susceptible. According to a standard continuous, deterministic SIR model approach (14), the effective reproduction number *R*_*t*_ = *R*_0_ ∗ *S/P* depends on the fraction of people still susceptible (*S/P*). Here we define *R*_0_ and *R*_*t*_ to explicitly exclude interventions such as social distancing or quarantine, because these are the interventions whose magnitudes are being optimized. However, *R*_*t*_ is intended to include fixed cost interventions such as improved ventilation.

Population-wide social distancing is parameterized as a value *D* that varies between 0 and 1 such that the reproductive number is proportional to (1 − *D*). I.e., a value of *D* = 0 indicates normal social contact, while *D* = 1 indicates complete isolation. Reductions in *D* come from a variety of behavior changes including working from home and reducing social contact, and also includes measures to mitigate the danger given contact, such as wearing masks, meeting outside and/or at greater physical distance.

Let *Q*_*i*_ denote the number of infectious people who are quarantined or isolated and *Q*_*n*_ denote the number of non-infectious people who are quarantined. The benefit from a targeted quarantine policy depends on 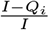, the fraction of infectious cases who are not successfully quarantined and then isolated. The effective reproduction number 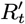 after both social distancing and quarantine interventions are applied is given by:

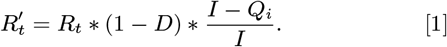

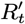 gives the expected number of onward transmissions per infected case in the general population. Equation 1 is undefined when the number of locally transmitted cases *I* = 0. Even for a low but non-zero value of *I*, a different, stochastic treatment warranted - this is discussed in Appendix B.

The per-individual cost of reduced contact is represented by the function *f* (*x*), where *x* is the individual counterpart to population-wide *D*, and where *f* (*x*) is assumed to be strictly increasing (from 0 cost with normal contact, to the maximum cost with complete isolation). The exact functional form of *f* (*x*) is unknown, but in Model Section B we show that the results have a bounded dependence on its form. The total cost to a population is given by Equation 2, based on *Q*_*i*_ + *Q*_*n*_ people in complete isolation, and *P* − *Q*_*i*_ − *Q*_*n*_ people with contact reduced by *D*.

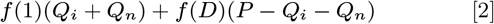

The amount of social distancing needed to meet a target reproductive number, *R*_*target*_, given fixed values for *Q*_*i*_ and *Q*_*n*_, is given by:

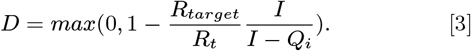

This value is bounded at 0; if 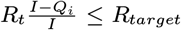, the target has already been met and no social distancing is required.

In the real world, it is not possible to adjust *D* instantaneously or exactly in response to changes in the impact of quarantine policy. However, in Appendix A, we show that Equation 3 is insensitive to time and control uncertainty, so long as caseload and control measures are fairly steady over time, as has been the case in many regions for at least some substantial period of time (8, 9).

Combining Equations 2 and 3, the total cost of a quarantine policy, in combination with a degree of population-wide distancing that keeps caseload steady, is given by:

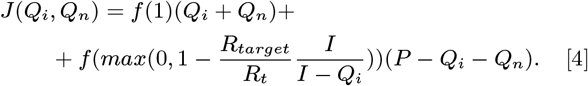

This equation can be used to compare the overall cost reduction from different quarantine policies.

We assume that an estimated risk of infectiousness is available when considering whether to recommend quarantine to an individual. Risk could be estimated using proximity and duration of contact with a known case as described in (12). Or it could be estimated for members of a subpopulation like a workplace by rapid testing of a random sample of that sub-population and projecting the proportion positive onto the remainder. When a person with risk of infectiousness *r* is quarantined, *Q*_*i*_ is incremented by 1 with probability *r*, and *Q*_*n*_ is incremented by 1 with probability 1 − *r*. Quarantining this person is worthwhile if the expected change in cost given by Equation 5 is less than 0:

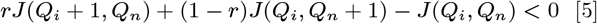

Solving the corresponding equation for *r*, the optimal risk threshold is given by:

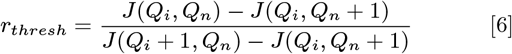

To apply Equation 6, we first consider the simplest equation for the cost of isolation, namely the linear function *f* (*x*) = *x*. Using this equation in a situation where some social distancing is necessary 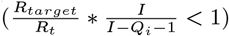, Equation 6 simplifies to:

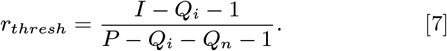

In this simple case, the interpretation of Equation 7 is straightforward and intuitive. The optimal risk threshold for quarantine is equal to the disease probability in the community, excluding from that community both those already isolated or quarantined, and the focal individual. This implies that in regions with very low prevalence rates, much stricter quarantine requirements should be used, making the general population less impacted by disease-control measures. In populations with higher disease prevalence in the community, there is less benefit to quarantining a single individual at low risk, because they represent a smaller fraction of the overall disease risk to the population.

### B. Sensitivity to Isolation Cost Function

We do not know the shape of the isolation cost function *f* (*x*) as the extent of isolation *x* varies. However, some non-linearity is expected. E.g., to achieve total rather than partial isolation can require a specialized quarantine facility in addition to a more extreme loss of individual utility. We define *D* to be constructed such that benefits in reducing disease transmission are linear in *D*. When considering a reduction in the extent of social activity, corresponding benefits are likely somewhere between linear and quadratic (5). When people seek out places that have variable evels of crowding, benefits are quadratic; when they seek out people who are still willing to meet with them, benefits are linear. By constructing *D* to have linear benefits, we pass this uncertainty in human behavior to the cost function. Disparate considerations all suggest a concave-up shape for *f* (*x*).

In Appendix C, we show that the deviation from Equation 7 is relatively small for reasonable choices of *f* (*x*). In particular, if the threshold *r*_1_ is computed using *f*_1_(*x*) = *x* and the threshold *r*_2_ is computed using any *f*_2_ (*x*) that is strictly increasing and concave up, then *r*_2_ ≥ *r*_1_ and 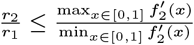. In practice, the value of *r*_2_ is often much closer to *r*_1_ than indicated by the upper bound. For example, while the ratio between optimal risk thresholds for the two cost functions shown in Figure 2 is bounded at a 10-fold difference, we numerically find it to be at most 4.1-fold.

**Fig. 2.**
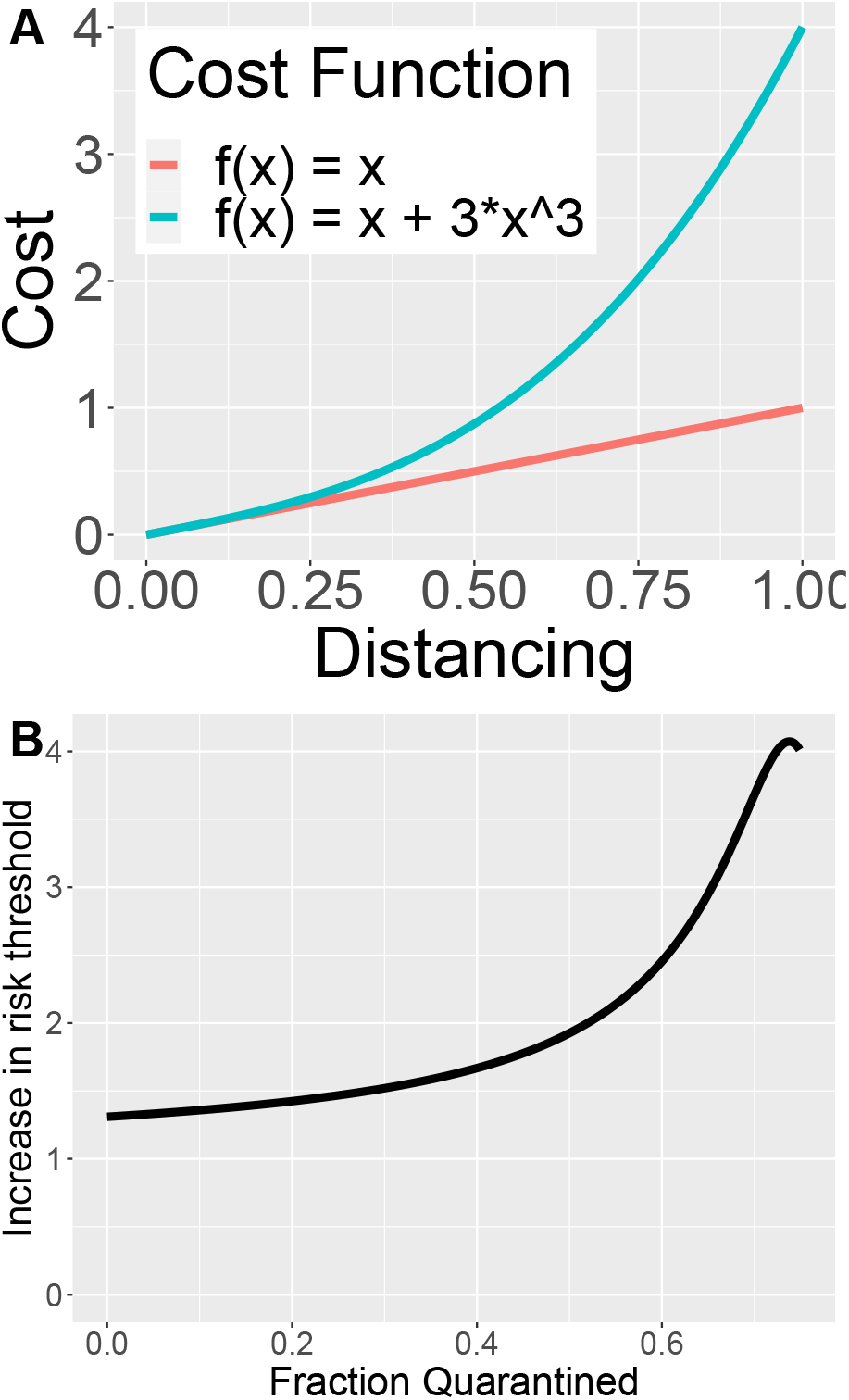
Non-linearity in the cost of isolation (A) leads to a modest four-fold change in optimal risk threshold, or less, depending for example on the proportion of infectious people quarantined (B).

Both the cost of isolation and the benefit of isolation can vary among individuals. For example, the cost of isolation is higher for essential workers than for those that can easily work from home, while the benefit of isolation is higher for those whose jobs expose more and/or more vulnerable people. Equation 7 can be modified to accommodate knowledge about an individual. If isolation is *m* times more expensive for this person than for the general population, and the potential danger to others if they are actually infectious is *d* times that of an average case, then the risk threshold for this person should be modified to *r*_*thresh*_ ∗ *m/d*. Note that this approach difers substantially from suggestions to target isolation directly to the elderly and other high medical risk individuals (5). We instead recommend increasing the stringency of quarantine among their contacts.

For a given individual, the cost of quarantine is likely to depend more than linearly on quarantine length, due both to logistical and to psychological factors. To accommodate this, the cost of quarantine could be modelled as time-dependent by scaling *f* (*x*) ∗ *m*(*t*). The benefit of quarantine is also not equal for each possible day of quarantine, given a probability distribution for incubation period and a quantitative timecourse of infectiousness (12). Consideration of these two factors will lead shorter recommended quarantine durations, targeted to the most infectious days. Testing individuals in quarantine could further shorten its duration (15).

The remarkable thing about Equation 7 is its insensitivity to *R*_*t*_ and *R*_*target*_. To determine the degree to which this might generalize, in Appendix B, we derive Equation 8:

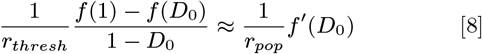

Equation 8 shows that *r*_*thresh*_ is chosen so that the risk-weighted cost of quarantine is equal to the marginal cost of distancing for the general population. Here, *r*_*thresh*_ depends indirectly on *R*_*t*_ and *R*_*target*_ because they influence the set point, *D*_0_. However, for a linear cost function, the slope (*f* ′ (*D*_0_)) and average slope 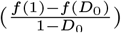 are constant, and therefore *r*_*thresh*_ only depends on the disease prevalence in the general population. In other words, when the cost per individual is a linear function of distancing, then the cost is the same whether that distancing is all directed at making one individual isolate completely instead of following current distancing norms, versus distributing the additional distancing among all individuals. For a non-linear function, these two costs are no longer equal. *D*_0_ is proportional to *R*_*target*_ */R*_*t*_, and so Equation 8 describes the degree to which non-linearity makes *r*_*thresh*_ depend on these terms. This dependence will be most pronounced when the local slope of *f* (*x*) around current distancing levels and the mean slope from current distancing to *D* = 1 are substantially different.

Because the optimal risk threshold isn’t very sensitive to the cost function, we focus our analysis on the more convenient linear cost function (with solution given by Equation 7).

## 4. Results

### A. Applications in time and place

In a large population with many cases, the risk threshold for quarantine *r*_*thresh*_ is closely related to the base rate of infection in the population. Here we give several examples in which we apply this finding, to illustrate how dramatically this causes optimal policy to vary across different places and times.

In British Columbia in October 2020, rapid SARS-CoV-2 testing was widespread, but only when symptoms were present, making official case counts of around 100/day a good estimate of symptomatic cases, such that total cases can be estimated in proportion. (When testing is less adequate, estimating the true number of cases is more complex, involving projections from death rates and/or testing rates, but has been attempted e.g. at (8) or (16).) Assuming another 20 asymptomatic or other undiscovered cases per day, each with a 10 day infectious window, yields 200 non-isolated infectious individuals on any given day. In addition, we estimate 4 infectious days per discovered case prior to isolation, yielding another 400 non-isolated infectious individuals on any given day. The base rate of infectiousness, conditional on not having been isolated, is thus of order 600/5,000,000=O(10^−4^). From Equation 7, a quarantine risk threshold of 10^−4^ is appropriate, or a little higher if a non-linear cost function is assumed in Equation 6. This implies that quarantine recommendations for entire schools or workplaces are worthwhile if they are expected to isolate one more infected person than would be achieved via traditional contact tracing.

In contrast, North Dakota in October 2020 had around 400 detected cases per day and perhaps another ∼400 undetected in a population of ∼800,000. This yields ∼5000 undetected infectious individuals on a given day, or a 0.6% base rate and corresponding risk threshold. Several studies have found the secondary attack rate to be on the order of 1/10 - 1/100 (17– 19), so this risk threshold aligns well with standard guidelines for the minimum contact considered to be close for contact tracing purposes. We note however that a 14 day quarantine is more than is needed to put the conditional probability of infectiousness, given lack of symptoms, of a non-household close contact below the base rate (12).

Imported and exported cases introduce complications. Policy is best set for regions that are relatively self-contained, and do not e.g. cut off commuter communities from one another. When imported and exported cases make up only a small fraction of total cases, then subtleties in using case counts to estimate 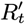 (discussed in Appendix B) have negligible effects. When local transmissions are abundant enough such that elimination is not a near-term goal, then incoming individuals can therefore be quarantined by comparing their infectiousness risk to *r*_*thresh*_ on the basis of base rates of infection in the location they arrived from, plus the risk of travel. These considerations apply e.g. to the need to quarantine following travel from one part of the U.S. to another.

Some locations, e.g. British Columbia, achieved very low levels of local transmission, but with a degree of economic connectedness with harder hit locations that made infeasible sufficiently strict quarantine to put the risk from incomers below *r*_*thresh*_ as described above (20). This is because the marginal cost of quarantine applies not just to the quarantined individuals, but to trucking routes and other aspects of the economic system. At this point, some significant expected number of imported cases acts as a forcing function outside the exponential dynamics of 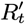. In this case, it might be better to set a target value for the expected number of cases per day, and then to set *R*_*target*_ to achieve this in combination with imported cases.

If local elimination is achieved, including successfully quarantining all imported cases, then *R*_*target*_ can be relaxed up to *R*_*t*_, removing all social distancing, with huge social gain. The disproportionate benefits warrant careful attention to avoid even a single imported non-quarantined case. How best to achieve this is best quantified by a stochastic model, outside the scope of this paper. Qualitatively, we note that the cost of letting a single case slip through, and hence having to return to social distancing, is higher for larger populations. The harm is also magnified by delays in realizing that an outbreak is underway, and hence the extent of harm depends strongly on local surveillance and contact tracing capabilities.

When different regions come under shared political control, importation risk can be controlled at the source rather than the destination. An example is Vietnam in March 2020. Viewed as an entire country instead of as smaller regions, it is optimal to quarantine at a threshold of 1*/*10^6^. This would motivate quarantining entire cities so that the rest of the country doesn’t have to substantially modify their behaviour. This is similar to the approach Vietnam actually took, with up to 80,000 people in quarantine at a time from regional lockdowns and aggressive contact tracing (21).

### B. Quantifying the Value of Quarantine

The value of a test, trace, and quarantine policy can be quantified in terms of the reduction it enables in full or partial person-days of isolation across a population. By definition, the marginal value of quarantining an individual of marginal risk *r*_*thresh*_ is 0. This implies, for a cost function that is uniform across individuals and neglects complications due to compliance, that quarantining one definitely infectious person for one day yields a benefit of *C/r*_*thresh*_ where e.g. *C* is the assessed cost of quarantine. Taking into account the quarantined individual, the net benefit is *C/*(1*/r*_*thresh*_ − 1).

In many countries, compliance not just with social distancing, but also with quarantine has been low. This is to be expected, given the uncompensated cost of quarantine to individuals, who are currently asked (or in cases coerced) to sacrifice for the public good. We advocate that governments align incentives, by guaranteeing e.g. 150% the individual’s normal daily income. This approach also serves to convert the costs and benefits calculated here, whose units are of person-days of isolation, to a dollar value for quarantine. On the basis of median income, and taking into account that some individuals are able to work from home and so will not draw the full 150%, this would in an affluent nation amount to something on the order of USD$150 per day of complete quarantine. This is the implied utility sacrificed by either total quarantine or equivalent social distancing *D*. I.e., a dollar value on the order of USD$150 per day is what one would need to pay people in order to incentivize their compliance, i.e. to achieve neutrality among preferences. Because this neutrality between quarantine plus payment vs. no payment and no quarantine does not include the dangers posed by COVID-19 itself, the existence of such payments would not incentivize deliberate exposure. Redistributive payments move costs from the quarantined individuals to the taxpayer.

With this cost applying to both indiscriminate distancing and targeted quarantining, we can assess the value of a quarantine policy from its ability to reduce the total amount of distancing required to achieve *R*_*target*_. Given the changing nature of a pandemic, we calculate how this total value depends on case prevalence and on the extent of population immunity.

When case prevalence is low, quarantining a single individual makes a larger proportional difference to the risk to others, enabling a greater relaxation of social distancing. We see in Figure 3B that in the pertinent range of 0.1%-5% case prevalence, the value of effective quarantine policies increases as case counts drop. Results are similar for the differently shaped distributions of assessable risk shown in Figure 3A. For lower case prevalence, the value flattens out because the benefit from quarantining one infectious person rises at the same rate that the number of total infectious people drops, while the number of infectious contacts quarantined per index case remains almost constant.

**Fig. 3.**
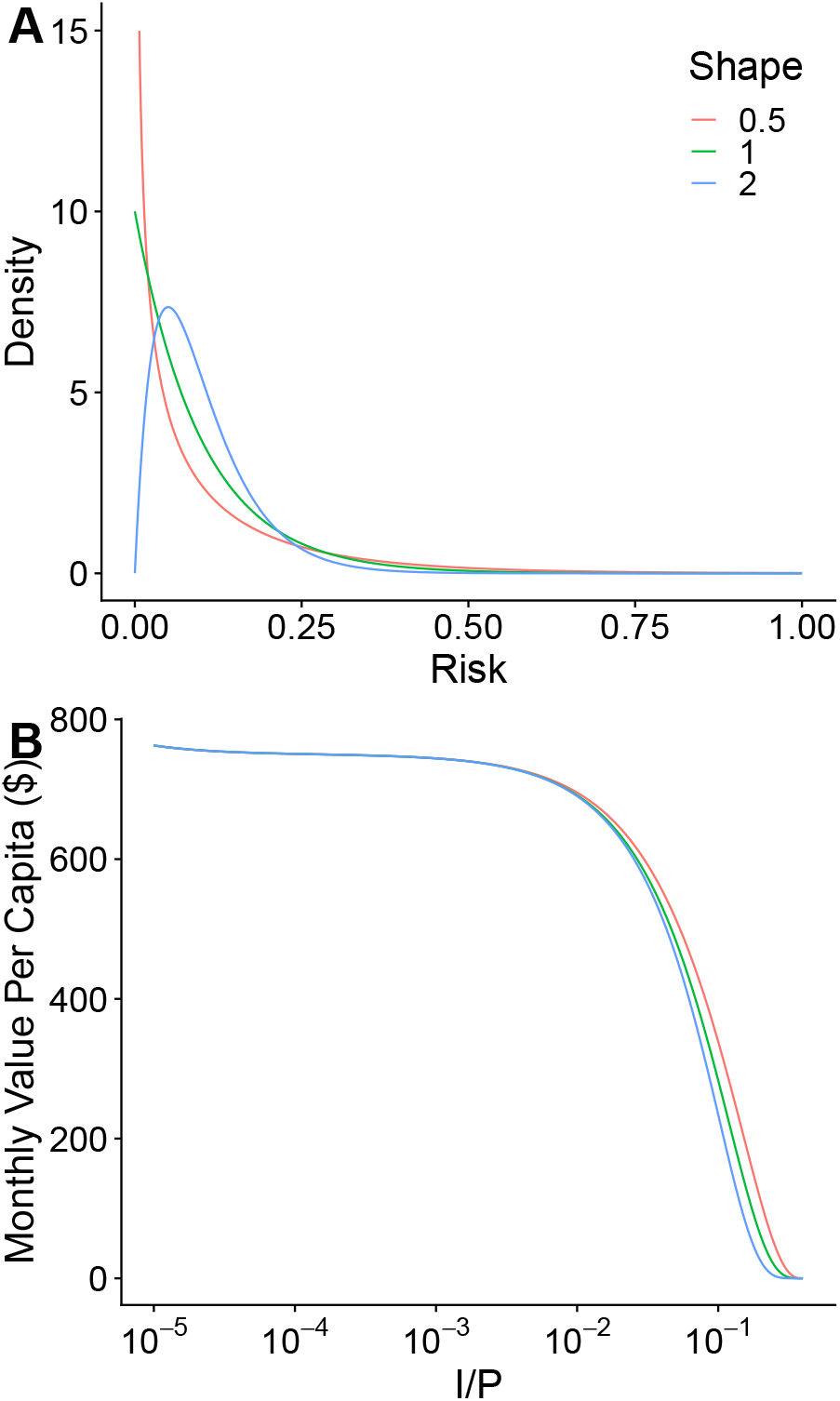
The benefits from quarantine policies are highest when case prevalence is low. In this example, each case one serial interval prior has on average ten contacts, of whom one is infected, such that 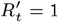. Higher case prevalence corresponds to high *r*_*thresh*_ and so lower *D*. As *D* decreases, the expected number of contacts each un-quarantined case has increases, but the number of positive cases in quarantine increases so that the overall expected number of contacts stays constant (maintaining 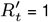). We assume that contact tracing succeeds in reaching 10% of all contacts. A) Assessable risk among contacts is modeled as a gamma distribution, in all cases with mean 0.1, and with different shape parameters to explore the importance of resolution among low-risk individuals. B) *Q*_*i*_ = 0.4*I, Q*_*n*_ = 4*Q*_*i*_, *C* = $150, *R*_*t*_ = 4, *R*_*target*_ = 1, *f* (*x*) = *x*. While *R*_*t*_ = 4 is higher than some estimates for SARS-CoV-2, other estimates place it as high as 5.7 even during early spread in China (22), and higher for new variants such as B.1.1.7.

Vaccination has enormous value in reducing the amount of distancing required to achieve *R*_*target*_. But unless the immune are exempted from distancing norms, the marginal value of quarantine, in helping sustain *R*_*target*_ with even less distancing, is unchanged. The effective reproductive number before including distancing, *R*_*t*_, is proportional to the susceptible fraction, *S/P*. Under a linear cost function, we see from Equation 7 that the threshold for quarantine is insensitive to *R*_*t*_. This means that the value of a quarantine policy is also insensitive to *R*_*t*_ and hence to this change in the vaccination status of the population. However, if people who have immunity are exempt from social distancing, not only is the amount of social distancing per person reduced, but also the number of people who have to distance. This has an additional impact on *r*_*thresh*_: the effective size of the population is reduced from P to S. Reducing the effective size of the population increases *r*_*thresh*_ by replacing *P* with *S* in the denominator of Equation 7.

In Figure 4, the value of a contact tracing system is shown as a function of the susceptible fraction of the population. If the immune are required to socially distance, the benefit of the program is independent of the proportion immune. If the immune are exempt, the benefit of the program is nearly proportional to the susceptible fraction. Intuitively, this is because quarantining infectious cases only allows an increase in social activity for the susceptible.

**Fig. 4.**
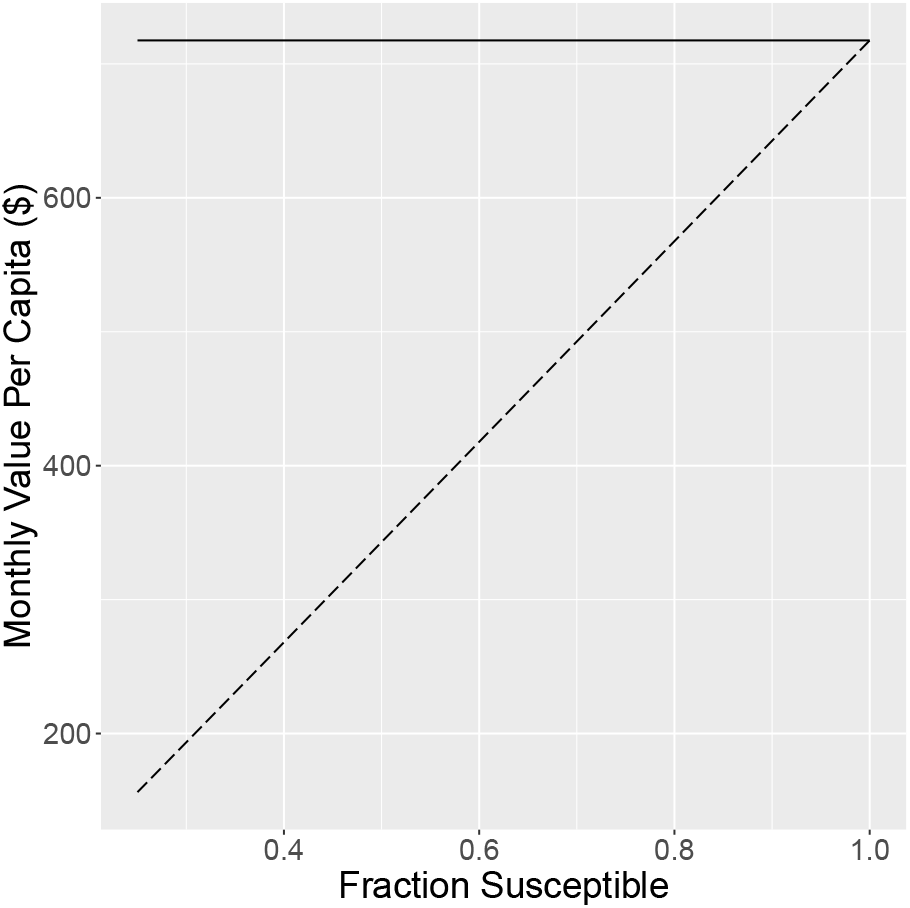
With vaccinated people exempt from distancing, the benefit of a contact tracing system is approximately proportional to the susceptible fraction (dashed line). When distancing is required, with the immune following the same distancing norms as the susceptible, the benefit does not depend on the susceptible fraction (solid line). *I/P* = 0.0055, *Q*_*i*_ = 0.4 * *I, Q*_*n*_ = 4 * *Q*_*i*_, *C* = $150, *R*_0_ = 4, *R*_*target*_ = 1 Assumptions about contact tracing are as for Figure 3. A linear cost function is shown, but the approximately proportional relationship also holds for a non-linear cost function, with immunity adjusting the effective population size.

### C. Marginal Value of Information

With a method available to assess the value of a quarantine policy, we can now assess the value of information in enabling better quarantine/isolation policies. We first consider random surveillance using a test with 50% sensitivity and near-100% specificity (the latter achieved via a follow-up PCR test). The value per test is given by the probability of testing positive * expected number of additional infectious days in isolation * daily benefit of each additional isolation day. If a positive test leads to 3 extra isolation days, and assuming a large enough population to neglect the − 1 term in Equation 7, the direct benefit (not including onward contact tracing) per test is 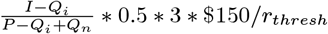. With 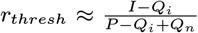, the per-test benefit is insensitive to the risk threshold and equal to ≈ $225. With a non-linear cost function, the benefit might be up to ten times less.

This makes random surveillance clearly worthwhile for rapid testing technologies that can be made available for as little as $5 per test (23). The benefit of surveillance testing increases with when close contacts are traced and quarantined. The benefit is reduced if a fraction of people who test positive do not effectively isolate. Note that previous estimates of the value of random surveillance supported less intensive measures (24), because they did not account for the benefits in terms of population-wide relaxation of other measures.

When testing is too expensive to be used for random surveillance, or merely too slow to roll out at sufficient scale, it can achieve higher marginal value if targeted to populations at higher risk. We note that it can be hard to justify strict quarantine on individuals of modestly above-average risk when the risk threshold is low in absolute terms, especially when elimination does not yet seem within reach. An alternative is daily testing instead of quarantine. The economic benefit from testing increases with risk of the focal individual relative to the average, and can thus justify more expensive and more sensitive daily tests.

### D. Distinguishing among Exposures

Current guidance from the World Health Organization (25) and the Center for Disease Control and Prevention (26) calls for quarantine for those within 1 meter or 6 feet, for 15 minutes, from 2 days before an infected individual’s symptom onset date to 9 days after. This guidance does not correspond to a consistent risk threshold, because it combines three binary thresholds rather than combining three risk factors prior to applying a threshold (12), because the infectiousness window does not reflect current science but instead a paper for which a formal correction has been issued (27–29), and because additional information such as ventilation in an indoor space is not taken into account. In addition, once a consistent risk assessment can be produced, we have shown here the benefits of adjusting the threshold based on local conditions.

While complex risk assessment can be difficult to apply in real-time case investigation interviews, exposure notification apps (30, 31) can easily do so (12, 32, 33). Here we consider the economic value of using a better rather than a worse risk assessment algorithm. Specifically, we consider the economic benefit that would arise from modifying the Google/Apple Exposure Notification protocol so that it could sense whether the user was indoor vs. outdoor, and differentiate risk between the two.

We assume a 50:50 distribution between indoor and outdoor exposures, each of which have a gamma distribution of risk with shape parameter 2 as in Figure 3, and a 15-fold difference in mean risk (34). We set the mean risk across both to be 0.06. When the risk threshold is low, there is no economic benefit from distinguishing indoor vs outdoor, for the simple reason that quarantine is recommended to anyone with any recorded exposure. However, when the risk threshold is high, knowing that an exposure occurred outdoors can spare an individual from costly quarantine, while also quarantining indoor exposures that occurred at greater distance.

With a 2% risk threshold, for the average exposed person, a fraction of whom are told to quarantine, the net benefit is 2.32 quarantine-days averted with the information and 2.05 without. These units of quarantine-days averted include both targeted quarantine and indiscriminate social distancing, which can be summed under a linear cost function.

Consider a population of 10 million, 2% of whom test positive over the time period of interest. We assume 20% use the app, of whom 50% enter their diagnosis into the app, thus notifying 20% of contacts that the app scores as above threshold risk, who go on to infect half as many people as they would were they not contacted. We assume the cost of quarantine is similarly only half what it would be given complete compliance. We assume that each has 16.7 contacts (set so that *R*_*t*_ = 1) following the distribution described above.

We assume that the cost of quarantine is $150 per day, and that the average quarantine is 10 days long. This gives an expected cost of quarantine of $1500 per individual that a particular configuration succeeds in getting to quarantine. At the margin, this indicates the value that society implicitly places, i.e. that removing a 0.02 risk is worth $1500. The benefit of quarantine is calculated in terms of “excess risk” above 0.02 having a value where each 0.02 of excess is worth $1500. Under these assumptions, net benefit comes out to $116 million with information about indoor/outdoor contact (i.e. $11.60 per capita or $58 per app user), and $105 million without. This value of $58 per app user can be used to calculate a marginal return on further investment in marketing for higher adoption, noting the non-linearity that will tend to produce accelerating rather than diminishing returns on investment.

We note that the value of an exposure notification app would be dramatically higher if, instead of a blanket 14 day quarantine duration, a shorter quarantine, targeted to the days of greatest risk, were used (12). Further gains would come from using negative test results to shorten quarantine.

## 5. Discussion

Here we showed that the socially optimal policy is to quarantine an individual if their risk of infectiousness is even mildly above that of the average person in the population who is not under quarantine. How much above depends slightly on the non-linearity of the cost of isolation with the strictness of isolation. While only order of magnitude calculations are sometimes possible, given that case prevalences vary over several orders of magnitudes across different populations, our rough calculations are nevertheless instructive. Some tools, like exposure notification apps, have quantitative sensitivities that can be tuned in real time in ways informed by such calculations. With more information about risk, quarantine policy can make a greater contribution to returning to a more normal life. The value of quarantine policy in doing so is higher for low case prevalence. It depends on vaccination prevalence only if the vaccinated follow different distancing norms than the unvaccinated, or if population immunity on its own is extensive enough to achieve *R*_*t*_ *<* 1.

To simplify the optimal trade-off between targeted quarantine and broad social distancing, we have neglected two complications. First, we have based recommendations as to who should quarantine on the assumption that they will do so completely. In reality, quarantined individuals often reduce rather than eliminate non-household contact, let alone all contact. More nuanced messaging regarding the degree of quarantine may be difficult to manage.

Second, how to choose *R*_*target*_ is not specified here. In the Introduction and Appendix A, we discuss a control theory perspective on the fact that social distancing adjusts to keep the long-term geometric mean of *R*_*t*_ near 1. A well-functioning control system is one with a negative feedback loop such that the error in achieving a desired outcome feeds in as a correcting input (35). Feedback can come either from top-down policy or from changes in individual decisions in response to conditions.

A control system can be described as a combination of proportional, integral, and derivative control (35). Derivative control would set social distancing policy based on whether exponential growth has resumed, and if so with what doubling time. This is unlikely to occur spontaneously, but rather requires government action in response to epidemiological reports not yet posing significant personal risk to the average individual. Spontaneous control by self-interested individuals responding to local conditions would under ideal circumstances follow proportional control, i.e. reflect current case counts. More likely, individuals will respond to the integral of case counts over time, e.g. as reflected in hospital occupancy rates. Targeting hospital occupancy rates (a combination of proportional and integral control) is also the explicit policy of some governments, e.g. the State of Arizona (36). If the goal is a managed approach to naturally acquired population immunity (37), hospital usage is a reasonable target. But since the amount of social distancing (whether mandated or voluntary) needed to keep *R*_*t*_ near 1 is the same against a background of low case counts as with high case counts, there are obvious health benefits to doing so with low case counts. Integral control will lead to significant fluctuations with a high total case burden.

We recommend mostly derivative control, with sufficient proportional control to ensure return to low case counts following fluctuations. The optimal extent of proportional control depends on whether shorter, sharper action vs. moderate decline with more social freedom achieves the least harm in transitioning between different case prevalences. The choice of *R*_*target*_ to control the speed with which case numbers are brought to low levels must also place a value on illness and death in comparison with social restrictions, a difficult problem.

Regardless of the form of feedback, compliance with transmission control measures such as size limits to gathering, compulsory masking etc. can be hard to predict, and hence the degree of top-down control is limited. This underlies our decision in this manuscript to focus on marginal costs given prevailing policy and behaviors. Our approach exploits the fact that some unspecified form of negative feedback keeps *R*_*t*_, or at least its geometric mean, near 1.

If testing and tracing improves, then less population-wide distancing will be required to achieve the same value of *R*_*target*_, and if the same level of distancing is maintaining, this will manifest as a drop in 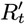. This might on occasion lead to a reconsideration of *R*_*target*_ to a lower level once that seems achievable at lower cost, but otherwise, reductions in social distancing should again ideally be triggered via derivative control, reducing the burden on the population.

While some form of negative feedback can be inferred from the empirical tendency of *R*_*t*_ to stay near 1, positive feedback can also occur: as case counts rise, contact tracing becomes more difficult (38), potentially accelerating the outbreak. Consciously adjusting the risk threshold that triggers quarantine, whether regarding incoming travellers, or risk thresholds in an exposure notification app, contributes to this positive feedback loop. However, conscious recognition of this positive feedback loop, and the limits it creates, can focus contact tracers’ attention where it can do the most good, in a manner that is under straightforward real-time control.

Because of the positive feedback associated with contact tracing workload and optimal quarantine policies, control of 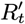 via negative feedback, e.g. to bring it back down to its target given a rise due to seasonal conditions or simply pandemic fatigue, is better exerted through population-wide measures to affect *D*. Our work shows that this is the socially optimal approach, and indeed if performed adequately, should be accompanied by a perhaps unintuitive relaxing of quarantine. Our formal development assumes a perfect control system, and focuses on one-individual perturbations to an equilibrium. It is likely that *D* will not reliably adjust in a completely timely manner, but a proactive approach can improve the chances of this occurring.

The value of quarantine policies is highest when case counts are low. However if low cases counts are a consequence of immunity (whether from natural infection or from vaccination), then the value of a quarantine policy will go down when those with immunity follow different distancing norms from those without. There is a direct benefit from relaxing distancing for the immune, whether this behavior is self-regulated or whether it comes in the form of a more official “immunity passport”. Immunity passports were once problematic because they would have set up an adverse incentive in favor of contracting SARS-CoV-2. However, with increasing vaccine availability, the benefits from immunity passports have risen at the same time as an adverse incentive to get infected has been converted into a pro-socially aligned incentive to get vaccinated. Our approach can be used to merge immunity history, quarantine policies, and even recent negative test results into an integrate metric of “risk to others”. The socially optimal policy is to use all available information to target restrictions to where they will do the most good, and restore as much social activity as possible while still controlling transmission.

## Data Availability

NA

## 6. Acknowledgements

James Petrie acknowledges support from the Ontario Graduate Scholarship and thanks his advisor, Prof. Stephen Vavasis, for giving him the freedom to work on this research project. We thank Amanda Wilson for helpful comments on an earlier draft, and the entire WeHealth team for their ethical commitment to maximizing benefits while minimizing harm, including via the recommendations of this paper. The opinions expressed in this publication are those of the authors and do not necessarily reflect those of WeHealth Solutions PBC.

### Appendices

#### A. Fluctuating 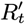

In Model Section A, we assumed that *D* instantaneously adapts to achieve *R*_*target*_ given prevailing conditions and current quarantine policies. This is motivated by the observation of relatively stable 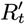 averaged over longer timescales. In this section, we show that Equation 1, and hence subsequent findings, are still a good approximation even when including stochastic effects, time delays, and uncertainty about system state.

To make this argument, we assume that for a certain region and time frame, the virus is neither eliminated nor creates herd immunity, and that control measures are relatively stable. Over *N* serial intervals between times *t*_0_ and *t*_*N*_, the initial and final case prevalence are related by 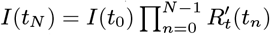. Equivalently, the geometric mean of 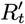 over this time period can be expressed using the initial and final case prevalence,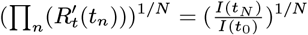. For most regions, case prevalence doesn’t vary by more than a couple orders of magnitude on the scale of months, which yields a (geometric) mean 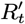 value quite close to 1. As an example, with *I*(*t*_*N*_)*/I*(*t*_1_) = 100, and *N* = 36 (6 months with a serial interval of 5 days), 100^1*/*36^ = 1.14. Approximating the geometric mean of the effective reproductive number as 1, and expanding to include distancing and quarantine terms:

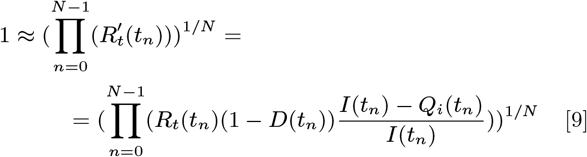

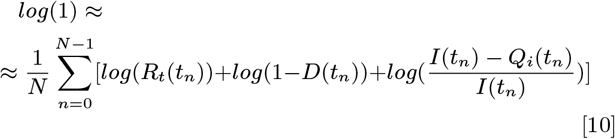

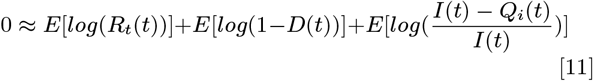

Assuming control measures don’t change drastically over time, we replace the mean of the log with the log of the mean:

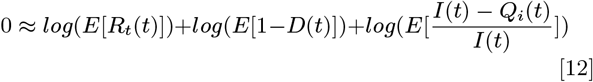

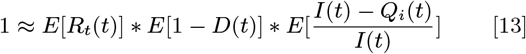

This is equivalent to Equation 1 with 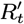 set to 1, showing that by averaging over time, the requirements for precise knowledge and control of system state can be relaxed.

#### B. Imported Cases and Stochasticity

For regions where imported cases make up a significant fraction of total cases, policy-makers should consider the modified dynamics when choosing *R*_*target*_. Imported cases act as a forcing term, with the number of cases at a future timepoint being the sum of locally obtained infections and imported infections. The mechanics of choosing a risk threshold do not change, but it is important to highlight that *R*_*target*_ only determines a portion of future cases. A complete solution must also consider the effect of border policies.

When the total number of cases is small, e.g. in a region with excellent local control for which rare imported cases are the primary concern, discrete and stochastic effects become important.

Quantitative models of tiny case count scenarios are challenging, because of stochasticity in the length of time needed to return to elimination, combined with curvature in the harms of shutdown as a function of its duration. This is further exacerbated by superspreader dynamics (39). Modelling the number of secondary cases with a negative binomial distribution with an overdispersion parameter 0.1 and mean 2.5 yields a variance of 65. In locations not aiming for local elimination, stochasticity averages out over time, and expected values can be used. When local elimination is being attempted, it would be useful to make use of discrete, stochastic models when choosing *R*_*target*_, e.g. to calculate the probability distribution of time to elimination and the expected damage from shutdowns of different severity. With the choice of *R*_*target*_ set, the previous arguments mostly still hold (although some consideration may be given to the difference in variance in transmissions based on broad vs. targeted distancing).

If local elimination has already been achieved, then a still more complicated case is to consider the risk of an outbreak from an imported case, the time delay and growth in the outbreak before it is noticed, and the subsequent shutdowns needed to re-achieve local elimination and thus relax general social distancing. It is likely that the considerable costs of such an outbreak warrant strict quarantine for incoming individuals. A robust testing and contact tracing system capable of quickly containing any new outbreak is also key.

#### C. Cost Function Dependence

Consider the situation where *r*_1_ is calculated using *f*_1_(*x*) = *x*, and *r*_2_ is calculated using *f*_2_(*x*), where *f*_2_(*x*) is strictly increasing, concave up, and differentiable. For both of these calculations, define the distancing set point 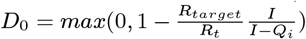, the required distancing if one more infectious person were isolated 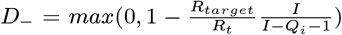, and *Q* = *Q*_*i*_ + *Q*_*n*_. By expanding Equation 6, the two risk thresholds are given by Equations 14 and 15:

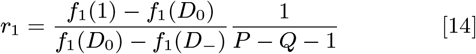

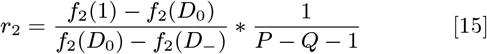

Substituting *f*_1_(*x*) = *x*, and taking the ratio of the two thresholds gives:

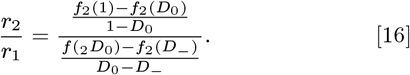

Since 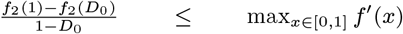, and 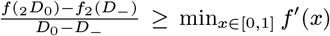, then Equation 17 gives a bound on the ratio of the two thresholds:

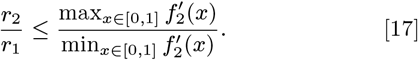

If *f*2(*x*) is concave up (indicating that additional isolation is more costly), then 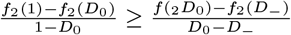 and therefore *r* _2_ ≥ *r* _1_.

Substituting Equation 7 into Equation 16, we obtain:

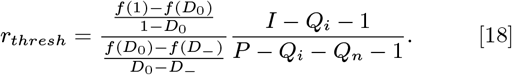

For differentiable *f* (*x*) and relatively large *I* − *Q*_*i*_, *D*_0_ − *D* _−_ is small, and 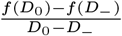 can be approximated as *f* ^!^(*D*_0_). Using the risk of the unquarantined population 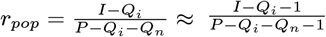, we obtain Equation 8.

